# A prospective study on tumour response assessment methods after neoadjuvant endocrine therapy in early oestrogen receptor positive breast cancer

**DOI:** 10.1101/2023.02.02.23285373

**Authors:** Joanna I. López-Velazco, Sara Manzano, María Otaño, Kepa Elorriaga, Núria Bultó, Julio Herrero, Ainhara Lahuerta, Virginia Segur, Isabel Álvarez-López, Maria M. Caffarel, Ander Urruticoechea

## Abstract

Neoadjuvant endocrine therapy (NET) in oestrogen receptor-positive/HER2-negative breast cancer (ER+/HER2-BC) allows real-time evaluation of drug efficacy and biological changes upon estrogenic deprivation. Clinical and pathological evaluation after NET may be used to obtain prognostic and predictive information of tumour response. Scales developed to evaluate response after neoadjuvant chemotherapy are not useful and there are not many validated biomarkers to assess response to NET. In this prospective study, we analysed radiological and pathological tumour response of 104 postmenopausal ER+/HER2-BC patients, treated with NET for a mean of 7 months. Our results show that radiological evaluation underestimates pathological tumour size, although it can be used to assess tumour response. In addition, we propose that the tumour cellularity size (TCS), calculated as the product of the residual tumour cellularity in the surgical specimen and the tumour pathological size, could become a new tool to standardize response assessment to NET. It is simple, reproducible and correlates with the existing biomarkers. Our findings shed light on the dynamics of NET response, challenge the paradigm of the ability of NET to decrease surgical volume and point to the utility of the TCS to quantify the scattered tumour response usually produced by endocrine therapy.

## Background

Oestrogen receptor positive (ER+)/human epidermal growth factor receptor 2 negative (HER2-) breast cancer (hereafter referred to as ER+ BC) represents almost 70% of all breast malignancies. Antiestrogenic or endocrine therapy is the cornerstone of ER+ BC treatment, being the neoadjuvant (preoperative) setting a very attractive scenario to find novel biomarkers of response and therapeutic strategies^1^. This is an urgent clinical need because long-term resistance to endocrine therapy is a common event^1^. Neoadjuvant endocrine therapy (NET) results in pathological and clinical response rates similar to those observed with neoadjuvant chemotherapy (NCT) although with lower toxicity^2,3^. Three pioneer clinical trials (IMPACT, PROACT and P024) demonstrated that NET is effective in downsizing ER+ BC and facilitating breast-conserving surgery (BCS) and showed greater efficacy for aromatase inhibitors compared with tamoxifen^4–6^. As a consequence of these and other studies, NET, given for 4–8 months, is nowadays recommended by international guidelines for postmenopausal women presenting ER+ BC^7–9^. An important advantage of NET is that it allows “in vivo” evaluation of response, hence granting real-time examination of drug efficacy as well as investigation of the biological and molecular changes that occur after estrogenic deprivation. However, the lack of useful biomarkers of long-term efficacy of therapy has precluded the development of the neoadjuvant strategy for endocrine therapies.

In the management of patients under neoadjuvant systemic therapy (either NET or NCT) two important evaluations are performed. First, a preoperative assessment of radiological tumour response (rad-TR) determines the response grade and establishes the surgical strategy^10–12^. Next, surgical specimens are histopathologically evaluated to obtain prognostic information according to pathological tumour response (path-TR) scales^10,12^. In the case of BC patients treated with NCT, there are well-stablished parameters to measure tumour response, such as RECIST criteria, Miller & Payne and Sataloff grading scales, and residual cancer burden value^13–16^. However, only Preoperative Endocrine Therapy Prognostic Index (PEPI) score and Ki67 levels have been validated as prognostic markers after NET^17–19^. Hence, tumours that show substantial down-staging after NET and present low Ki67 levels and PEPI score at surgery have an excellent long-term prognosis even without chemotherapy^1,11,19,20^. Both Ki67 levels after NET and PEPI score have been shown to predict long-term outcomes (e.g., relapse-free survival). However, they are not optimal and they are not routinely used due to, among other reasons, a lack of Ki67 measurement standardisation^21^. In the clinical practice, understanding the impact of tumour response to NET in long-term outcome will help clinicians to individualize adjuvant treatment for ER+ BC. In contrast to what happens for NCT, pathologic complete response (pCR) after NET is a rare event and is not a useful marker of prognosis given its low likelihood^3,17,22^. In fact, previous studies suggest that ER+ BC tumours after neoadjuvant systemic therapy present a “diffuse cell loss” response at pathological level, which is characterized by a distribution of the tumour in multiple scattered foci or small groups of tumour cells without affecting overall tumour size^23^.

In this context, there is an urgent need for the identification of robust, reproducible biomarkers of response to NET with long term prognostic value. Ideally, these new biomarkers should be candidates for initial validation in retrospective series. In addition, the mentioned diffuse cell loss in ER+ BC patients treated with NET needs to be better characterized. In order to investigate the dynamics of tumour response, we generated a prospectively collected series of ER+ BC patients treated with NET. We characterised and compared tumour response by ultrasound scan (USS) and/or magnetic resonance imaging (MRI) with pathological tumour size (path-TS). Finally, we described a new biomarker with potential prognostic implications, called tumour cellularity size (TCS), which could help to characterize the response to NET in ER+ BC by an estimation of the diffuse cell loss.

## Methods

### Study population

We analysed clinical data from a cohort of patients treated in our institution between 2005 and 2019 following a homogenous therapeutic protocol. Data were prospectively collected and retrospectively analysed. All were postmenopausal women with histologically confirmed, untreated, invasive, operable, larger than 10 mm and amenable for radiological follow-up, ER+/HER2-non-metastatic breast cancer. Patients had to be treated for at least 3 months with NET prior to surgery with curative intention. The administered NET was an aromatase inhibitor except contraindicated. Informed consent was obtained from all patients.

### Imaging and histopathological analysis

Tumour baseline assessment was performed by breast USS and/or MRI. Clinical response by USS was evaluated after 2 months of treatment and repeated every 2 months. MRI and/or USS were also performed before surgery to evaluate radiological tumour response (rad-TR). Surgical breast specimens were evaluated by the pathologist to determine pathological tumour response (path-TR), tumour size (path-TS) and residual tumour cellularity (%). Clinical (assessed by MRI and USS) or pathological tumour size corresponds to the major diameter of the tumour in millimetres (mm) and T-stage of the primary tumour was defined according to AJCC Cancer Staging Manual^24^.

Immunohistochemistry analyses were performed in baseline formalin-fixed paraffin-embedded biopsies and surgical specimens to determine the expression of ER, progesterone receptor (PgR) and Ki67 levels, using international standards^25,26^. ER, PgR and Ki67 were recorded as continuous variables. The positivity cut-off for ER and PgR was ≥10% of stained nuclei. Ki67 score was defined as the percentage of tumour cells with Ki67 positive nuclear staining. At least 1000 tumour nuclei were counted per sample, according to the recommendations of Dowsett et al., 2011^25^. The change in Ki67 (ΔKi67) after NET was calculated using the following equation: ΔKi67 = [(Ki67 (%) in surgery specimen) – (Ki67 (%) in baseline biopsy)] / (Ki67 (%) in baseline biopsy). Consequently, ΔKi67 values ranged between -1 to 1. ΔKi67 results were categorized into three groups depending on their magnitude of change. ΔKi67 = -1 means that Ki67 changes to zero in the surgery specimen. ΔKi67= > -1 to < 0 means that the tumour presented a decrease in Ki67 expression. Finally, ΔKi67 ≥ 0 means that the tumour did not present any change in Ki67 expression or that Ki67 expression in the surgery specimen was greater than the one on baseline biopsy.

Rad-TR was defined using mRECIST 1.1 criteria^13^. According with this criteria, complete responses (CR) were defined as tumour disappearance and partial responses (PR) were defined as the reduction of the tumour diameter by ≥ 30%. An increase ≥ 20% in tumour diameter was qualified as progressive disease (PD). The rest of situations were qualified as stable disease (SD).

Path-TR was quantified using a modified Miller and Payne grading scale^14,27^. In this scale, response grades 1 and 2 (no change or less than 30% loss of tumour cells, respectively) were regarded as SD. Grades 3 and 4 (reduction in tumour cells between 30-90% and > 90%, respectively) were considered as pathologic PR. Grade 5 (defined as no malignant cells identifiable in the tumour niche) was considered pathologic CR (pCR). In binary analyses, path-TR was defined as loss of tumour cells ≥ 30 % (grades 3-5) and no path-TR as <30% (grades 1-2).

Modified PEPI (mPEPI) score was determined on the basis of tumour characteristics of surgical specimen (i.e. tumour size, nodal involvement status and Ki67 staining), as previously published^18,28^. Patients were classified into 3 mPEPI risk groups (I=0, II=1-3 and III=4+).

TCS, the novel score we introduce in this study, was calculated as the product of tumour cellularity in the surgical sample (%) and tumour diameter (path-TS, in mm).

### Statistical analyses

Statistical analyses were performed using GraphPad Prism version 9. For the descriptive statistical analyses, minimum, maximum and mean values were used. For Gaussians distributions, paired Student’s t-test was used to compare differences between two groups. For non-Gaussian distributions, Wilcoxon matched-pairs or Kruskal-Wallis tests were performed. Chi-square or Fisheŕs tests were used to determine differences between expected frequencies. Spearmańs r coefficient (rho) for analyses, were used to quantify correlations (both with a 95% of confidence interval). *P* values < 0.05 were considered statistically significant. Unless otherwise specified, histograms represent mean values +/- standard error of the mean (SEM).

## Results

### Tumour characteristics and change in tumour biomarkers after NET

104 patients with early ER+/HER2 breast cancer were included in our study. The study population presented a mean age at diagnosis of 69 (47-93) years and the mean NET duration before surgery was 7 months (3-39). The mean tumour size was 25 mm (10-90) assessed by MRI and 18 mm (40-70) by USS. The main administered NET drug was letrozole (n=100), but some patients also received anastrozole (n=2), tamoxifen (n=1), or exemestane (n=1). One patient was diagnosed with bilateral disease and her two tumours were independently considered in our analyses. The principal characteristics of the tumours and their surgical management as well as the pathological changes after NET are summarised in Table 1. No significant decrease of histological grade was observed after NET (*p* = 0.12). Ki67, ER and PgR expression was assessed in all tumours pre- and post-NET treatment. As seen in previous studies using similar cohorts (Toi et al. 2011; Martí et al. 2022), NET significantly decreased all these three parameters, being the changes in Ki67 and PgR the most significant (both *p* < 0.0001, Table 1 and supplementary material, Figure S1). While only 13 patients (12%) were cN+ before treatment, 26 patients (25%) were pN+ at pathological assessment. Regarding pathological tumour response to NET, only one case of pCR was recorded and most cases (72%) showed partial path-TR (Table 2). Most patients (81%) fell into low (I, n=34, 35%) and intermediate (II, n=45, 46%) mPEPI risk groups. These results are in agreement with the response rates obtained in similar series^8,10,12^.

**Table 1.** Histopathological information and surgical management of tumours included in our series. *One patient was diagnosed with bilateral disease and her two tumours were independently considered in the histopathological analysis and in its surgical management. ªOne tumour was not evaluable for biological characteristics at surgery because the patient achieved a pCR. c/yp axillary node status was determined clinically and pathologically before and after NET, respectively. BCS: Breast-conserving surgery. N/A: not available.

| Characteristics | Before NET<br>(n = 105*) | After NET<br>(n = 104<sup>a</sup>) |
| --- | --- | --- |
| Histological grade [n (%)] |  |  |
| I | 23 (22) | 34 (33) |
| II | 78 (74) | 69 (66) |
| III | 4 (4) | 1 (1) |
| Histological type [n (%)] |  |  |
| Ductal | 86 (82) | 85 (82) |
| Lobular | 10 (10) | 12 (12) |
| Other special type | 9 (8) | 7 (6) |
| c/yp axillary node status [n (%)] |  |  |
| Negative | 91 (87) | 72 (69) |
| Positive | 13 (12) | 26 (25) |
| N/A | 1 (1) | 7 (6) |
| Positive cells (%) [mean (range)] |  |  |
| Ki67 | 20 (3-60) | 9 (0-75) |
| Oestrogen receptor | 94 (20-100) | 90 (0-99) |
| Progesterone receptor | 63 (1-100) | 15 (0-99) |
| Breast surgery performed [n (%)]* |  |  |
| BCS |  | 95 (90) |
| Modified radical mastectomy |  | 10 (10) |

**Table 2.** Radiological (Rad-) and pathological (Path-) tumour response (TR) after NET. Rad-TR was evaluated by mRECIST 1.1 criteria and path-TR was measured using a modified Miller and Payne grading scale. N/A: not available.

| Rad-TR type [n (%)] | MRI | USS | Path-TR [n (%)] |
| --- | --- | --- | --- |
| N/A | 7 (7) | 14 (13) | 0 (0) |
| Complete | 27 (26) | 16 (15) | 1 (1) |
| Partial | 40 (38) | 49 (47) | 76 (72) |
| Stable disease | 29 (27) | 24 (23) | 28 (27) |
| Progressive disease | 2 (2) | 2 (2) | 0 (0) |

### Radiological examination of tumour size after NET underestimates pathological tumour size

To determine which is the best radiological technique to predict pathological tumour size (path-TS) after NET, we compared tumour size measured by MRI and USS before and after treatment. As expected, radiological tumour size (rad-TS), measured by MRI or USS, both at diagnosis and after NET (just before surgery), significantly correlated with path-TS (Figure 1A-D). Surprisingly, our results showed that path-TS correlated better with tumour size assessed by MRI and USS at diagnosis than after NET (Figure 1A). This may suggest that the radiological evaluation before surgery may not be a very precise technique to assess tumour size after NET. To better visualize this, we compared the mean value of tumour size assessed by each radiological technique, before and after NET, and by path-TS. As shown in Figure 1E, MRI/USS measurements after NET were significantly lower than path-TS and, interestingly, radiological measures at diagnosis were more similar to path-TS than the measures after treatment. Actually, MRI and USS before surgery underestimated path-TS in 77% (76/99) and 92% (84/91) of the cases, respectively.

**Figure 1.**
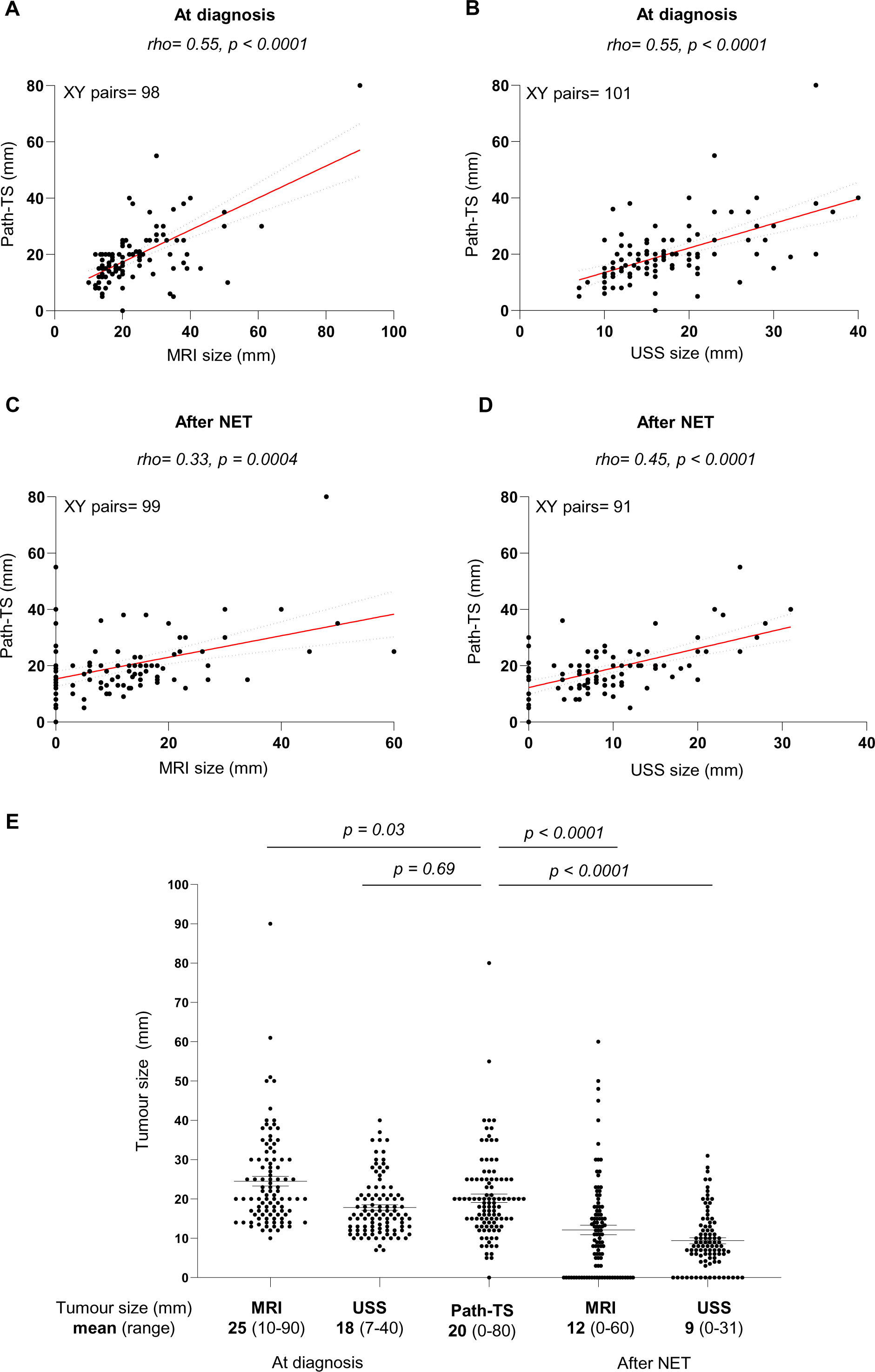
Comparison between pathological and radiological tumour size before and after neoadyuvant endocrine treatment (NET). (A-D) Correlation of pathological tumour size (path-TS) with MRI (A and C) and USS (B and D) measurements at diagnosis (A-B) and after NET (C-D). Spearman correlation coefficients (rho) and *p* values are shown. (E) Comparison of radiological tumour size (assessed by MRI and USS at diagnosis and after NET) with path-TS. *p* values were calculated using Kruskal-Wallis test.

Importantly, we also found that this disagreement in tumour size estimation by imaging and histopathological analysis affects the concordance between radiological (rad-TR) and pathological (path-TR) tumour response (Table 2). Complete rad-TR was observed in 27 (by MRI) and 16 (by USS) patients while only one patient presented a pCR by pathological assessment. To better visualize these discrepancies, we plotted the correlation between rad- and path-TR. As shown in Figure 2A-B, we found that rad-TR assessed by MRI correlated better with path-TR than rad-TR assessed by USS, although both associations were statistically significant. Interestingly, we observed that a considerable number of tumours presented a complete (100%) rad-TR after NET but presented a low path-TR (G2 or G3, highlighted in red in Figure 2A-B). Taken together, our data indicate that the radiological examination of tumour size after NET and before surgery underestimates path-TS bearing surgical implications for the definition of tumour area.

**Figure 2.**
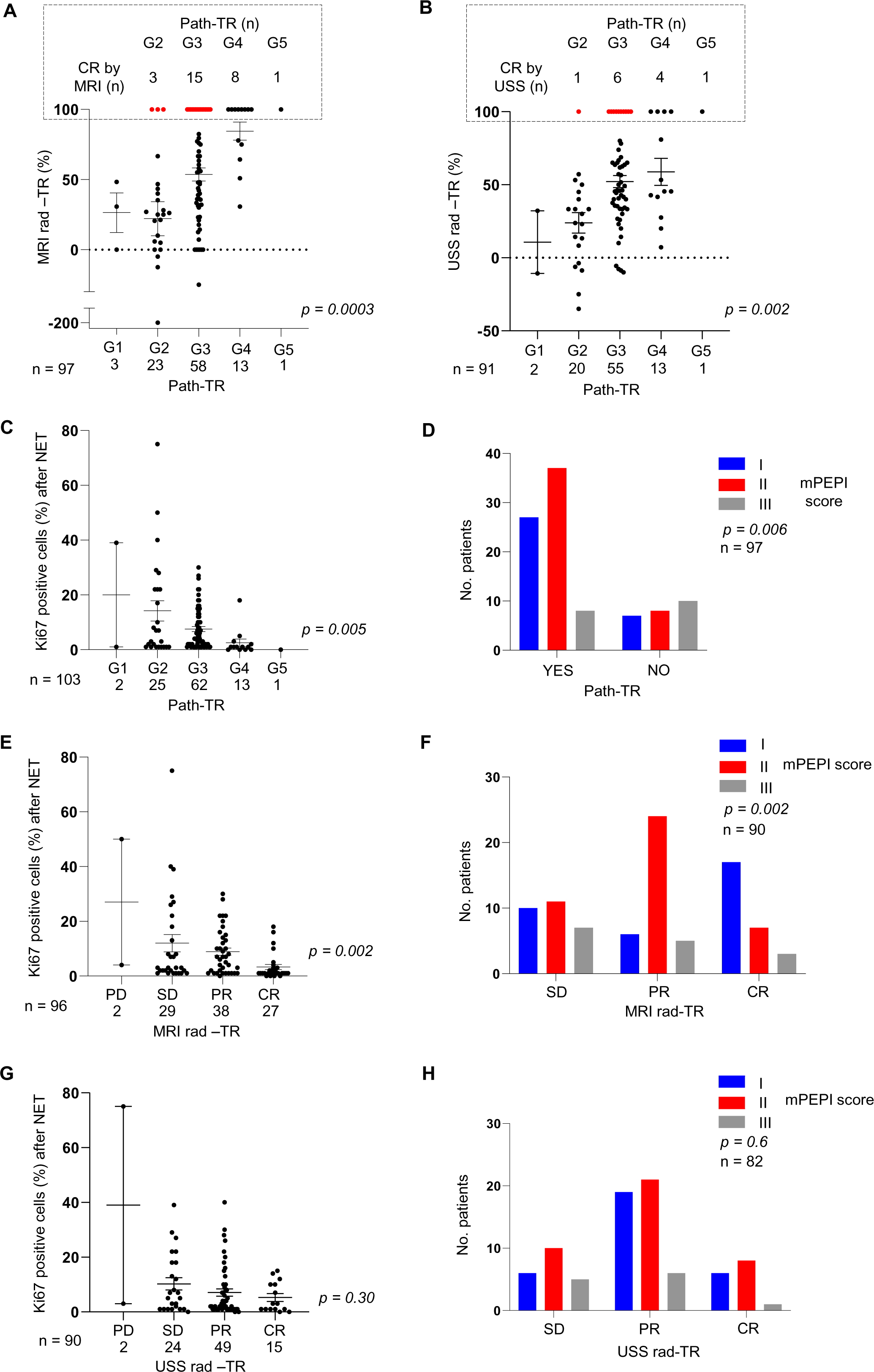
Evaluation of radiological tumour response (rad-TR) after NET by comparison with pathological tumour response (path-TR) and prognostic biomarkers for NET. (A-B) Rad-TR was assessed by MRI (A) and USS (B) and evaluated by mRECIST 1.1 criteria. Path-TR was evaluated using a modified Miller and Payne grading scale. In the upper square, tumours presenting complete response by MRI (A) or USS (B), but partial response (G2-G3) by path-TR, are highlighted in red. (C, E and G) Analysis of Ki67 levels at surgery in tumours classified according to thei*p* values were calculated (*All*). (D, F and H) Contingency analyses of the association between modified PEPI (mPEPI) score and path-TR (D) and rad-TR by MRI (F) or USS (H). *p* values were calculated using Kruskal-Wallis test (A-C, E, G) or Chi-square test (D, F and H). CR: complete response, PR: partial response, SD: stable disease, and PD: progression disease.

Next, we evaluated the association between rad- and path-TR with the two most accepted prognostic markers after NET: Ki67 levels and mPEPI score^11,18,28^. As expected, pathological responders presented significantly lower Ki67 levels at surgery and mPEPI score (Figure 2C-D). Regarding rad-TR, both prognostic markers were associated with tumour response assessed by MRI (Figure 2E-F), but, in the case of USS, there was not association between tumour response and Ki67 and PEPI score (Figure 2G-H).

In summary, our data support that radiological evaluation of tumour size after NET underestimates pathological tumour size and indicate that MRI could be more reliable than USS to assess response to NET.

### Tumour cellularity size is a new parameter to standardize the assessment of residual tumour content after NET

Diffuse cell loss has been observed as a common pattern of tumour response after neoadjuvant therapies in ER+ (luminal) tumours^23^. In an attempt to better assess tumour response after NET, we propose a novel parameter called tumour cellularity size (TCS). TCS is the product of tumour cellularity (%) and tumour diameter (path-TS, in mm) and estimates the volume of remaining cells in the tumour bed after NET. First, we evaluated how TCS relates to radiological tumour size and response (Figure 3). As seen in Figure 3A, TCS values were much lower than path-TS and more similar to MRI or USS measures after NET compared to rad-TS at diagnosis or path-TS. We then analysed how TCS associates with radiological and pathological response (Figure 3B-D). Our results showed that TCS inversely correlated with path-TR and with MRI rad-TR (Figure 3B-C). However, the association between TCS and rad-TR determined by USS was not significant (Figure 3D), in line with previous results supporting that MRI may be more adequate than USS to quantify response to NET. Taken together, our data indicate that TCS can quantify the tumour ‘’diffuse cell loss’’ response observed in ER+ BC tumours after NET, and may capture better the biological response of those tumours and explain why the radiological pre-operative assessment of tumour size underestimates the path-TS.

**Figure 3.**
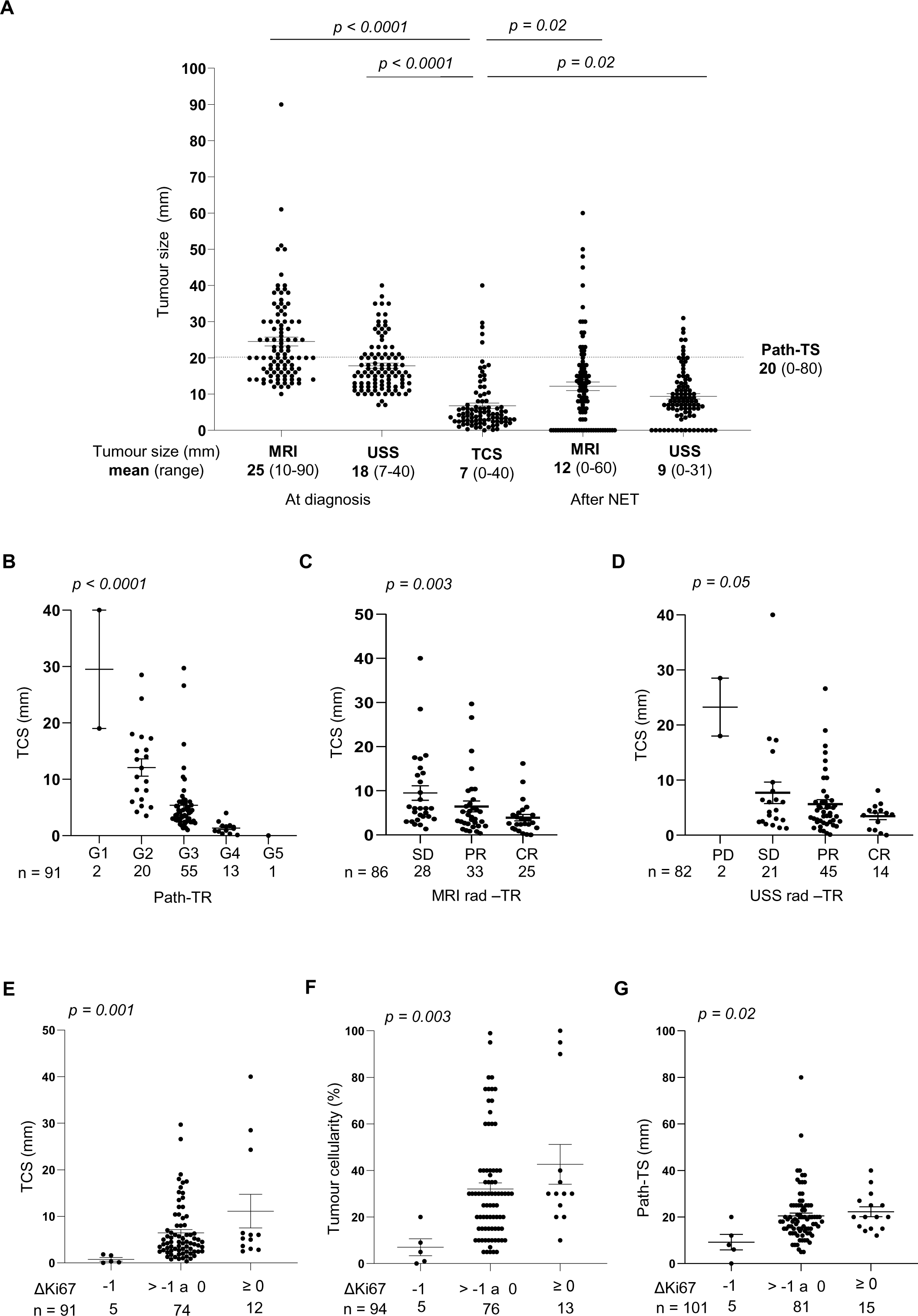
Tumour cellularity size (TCS) as a new parameter to measure response to NET. (A) Comparison of radiological tumour size (assessed by MRI and USS, before and after NET) with TCS. Dotted line indicates the mean of path-TS obtained for those tumours. (B-D) Association between TCS and pathological (path-TR) (B) and radiological response assessed by MRI (C) or USS (D). (E-G) Association between TCS (E), tumour cellularity (F) and pathological tumour size (path-TS) (G) with changes in Ki67 (ΔKi67) after NET. *p* values were calculated using Kruskal-Wallis test.

In order to further evaluate if TCS can be used as a biomarker of response and prognosis for patients undergoing NET, we evaluated its association with changes in Ki67 (ΔKi67) (Figure 3E-G) and Ki67 levels at surgery (supplementary material, Figure S2), well-established prognostic markers after NET. We observed that ΔKi67 and Ki67 expression at surgery correlated better with TCS than with tumour cellularity or path-TS (Figure 3E-G, and supplementary material, Figure S2A-C). Consequently, tumours with high residual Ki67 expression (ΔKi67 > 0 and high Ki67 expression at surgery) also present a high TCS, suggesting that TCS could be a promising biomarker of response to NET.

Finally, to identify an initial cut-off value for which TCS can divide patients responding to NET from no responder patients, we analysed the relationship of TCS quartiles with Ki67 (Figure 4 and supplementary material, Figure S3A-B). As mentioned before, TCS is positively correlated with Ki67 at surgery and ΔKi67 (Figure S3). Tumours with TCS <2.5 mm (Q1) showed significantly lower Ki67 levels at surgery and ΔKi67 compared with tumours with TCS ≥ 2.5 mm (Q2, Q3 and Q4, Figure 4), suggesting that a TCS value <2.5 mm could be used as a good cut-off value to identify patients responding to NET. However, more studies in independent cohorts with associated survival data are needed.

**Figure 4.**
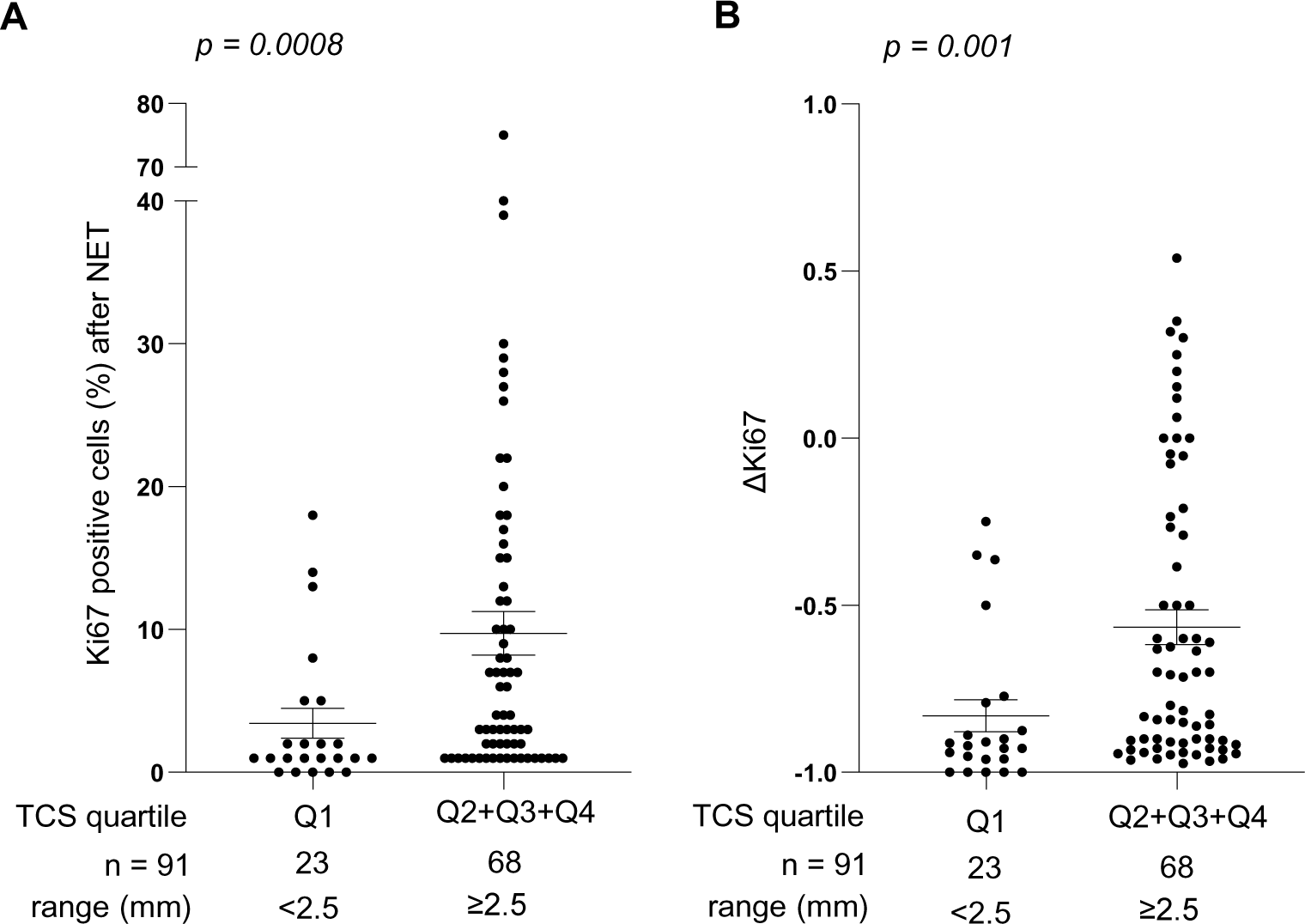
Identification of a tumour cellularity size (TCS) cut-off value to discriminate patients according to their response to NET. (A-B) Ki67 levels at surgery (A) and ΔKi67 (B) in tumours with low (quartile 1, Q1) versus high (Q2, Q3 and Q4) TCS. *p* values were calculated using Mann Whitney test.

## Discussion

There are different reasons why neadjuvant endocrine therapy (NET) is a very promising and attractive therapeutic strategy for ER+ BC patients. First, it is less toxic than neoadjuvant chemotherapy albeit resulting in similar pathological and clinical response rates and, indeed, it is already recommended by international guidelines for post-menopausal women^3^. Finally, it represents an ideal scenario for clinical research as it allows real time investigation of drug efficacy and of the molecular and biological changes in tumours after endocrine treatment. This may lead to the identification of novel biomarkers of response and new therapeutic strategies. However, NET remains an underused tool for ER+ BC because monitoring response is challenging, among other reasons^3,29^. Many NET clinical trials use the radiological response rate (by pre-operative evaluation with USS, mammography or MRI using RECIST 1.1 criteria) and improvement of BCS rates as a primary objective to demonstrate effectiveness^3,11,12^. However, our results, obtained from a prospectively collected series of 104 ER+ BC patients treated with NET, prove that the pre-operative radiological evaluation after NET underestimates path-TS. Previous reports have also shown that radiological and clinical evaluation after NET underestimate the lesion size^10^, although Reis and cols found this difference negligible^30^. Clinically, these discrepancies between radiological and pathological responses may have strong surgical implications for the definition of lesion area and our data suggest that the radiological evaluation of tumour size should not be determinant to plan the resection area. In fact, the AJCC recommends that imaging findings after NET, NCT and radiotherapy are not considered elements of initial clinical staging^24^.

In addition, our data show that radiological complete responses almost never parallel pCR. In our series, 26% and 15% of patients showed complete radiological response by MRI and USS, respectively, but only 1 patient achieved pCR. In fact, pCR after NET is a rare event and only occurs in less than 1% of the cases^3^. Usually, residual disease is found even in very good responder tumours, in the form of microscopically scattered residual cancer nests in the tumour bed [30]. This scattered or diffuse cell loss response is also observed after neoadjuvant chemotherapy in ER+ tumours^23^.

While pathological assessment considers the maximum area occupied by the tumour and does not capture this scattered response, radiological evaluation after NET may reflect the diffuse cell loss response. This could explain why rad-TS after NET did not reflect path-TS in the surgery specimens in our cohort and challenges the paradigm of NET as a tool to increase the BCS rates for ER+ BC. The difference between radiological and pathological evaluation of neoadjuvant systemic treatment (NST) is less frequently observed in triple-negative and non-luminal HER2+ tumours, which tend to present a shrinkage or concentric response (also called tumour collapse) to NST^23^.

Nevertheless, we should also take into consideration that the complete response assessed by MRI may capture biological events with potential prognostic/predictive value, including normalization of tumour vessels. Hence, the event of CR by MRI, even without pCR, may define a prognostic category that deserves further study.

Despite the discrepancies between radiological and pathological evaluation of tumour response to NET, clinical and radiological monitoring of tumour response during the course of NET are necessary to early detect disease progression. Our data indicate that MRI is preferable than USS to assess response to NET, as 1) path-TR correlated better with rad-TR assessed by MRI than by USS and 2) MRI rad-TR was significantly associated with reduction of Ki67 in the surgical specimen and lower mPEPI score, the two most accepted prognostic markers after NET^11,18,28^. This association was not statistically significant in the case of USS. Our results are in agreement with previous reports supporting the use of MRI as the most accurate tool among other methods (clinical examination, USS and mammography) to assess tumour response to NST in breast cancer^3,31^.

As mentioned above, the diffuse cell loss response observed in ER+ tumours as a response to NST represents a challenge to evaluate tumour response to NET. We hypothesize that the parameter tumour cellularity size (TCS), presented herein, can be used to assess the scattered response to NET as it estimates the multiple scattered foci of tumour cells in the tumour niche. TCS is the product of tumour cellularity (%) and tumour diameter (path-TS, in mm) in the post-treatment surgical sample. We found that TCS significantly associated with rad-TR evaluated with MRI. As previously discussed, only PEPI score and Ki67 expression under treatment are validated prognostic markers for NET^17–19^. Importantly, TCS correlated better with ΔKi67 than the percentage of tumour cellularity in the post-treatment sample and the path-TS. This may indicate that reduction in Ki67 expression is related to the tumour cellularity content even when the path-TS does not change after NET. The association between TCS and mPEPI score could not be evaluated as they are not independent variables since both include path-TS in their calculation^28^.

In summary, our results shed light on two clinically relevant and unanswered questions in the context of NET highlighted by Sella and cols^3^. One of them is which is the optimal imaging technique to pre-surgically evaluate residual disease after NET. Our data supports the use of MRI over USS, but also prove that both imaging techniques underestimate pathological tumour size. As mentioned, this points to careful consideration of clinical and radiological TR to define the surgical resection tumour area and challenge the paradigm of the reduction of surgical volume by NET given that the initial radiological assessment seems to be the best value to define the tumour area even after therapy. The second unanswered question is the need of novel biomarkers to assess pathological response to NET. We propose a new biomarker called tumour cellularity size (TCS) that could be a promising candidate to use in combination with changes in Ki67, although it should be validated in larger cohorts.

## Additional information

Supplementary information is available at BJC’s website

## Acknowledgements

We are grateful to the members of our laboratory for critical discussion of this work and to the Pathology Services of Hospital Donostia and Onkologikoa Foundation for technical assistance.

## Ethics Approval

The study was conducted in accordance with the Declaration of Helsinki and Good Clinical Practice guidelines as well as authorised by the Spain Health Authority and the local Ethics Committee.

## Authors’ contributions

JILV, SM, AU and MMC analysed the data and wrote the manuscript. AU designed the study. AU and MMC co-supervised the study. KE analysed histopathology. VS and NB analysed imaging data. JILV, MO, AL, JH, IAL and AU collected and analysed patient data.

## Funding

This work was funded by Instituto de Salud Carlos III (ISCIII) grants: PI21/01208, PI20/01253, CP18/00076 and FI19/00193 co-funded by the European Union, Basque Department of Health (2020111040), Fundación SEOM (SEOM Avon Fellowship 2020) and Ikerbasque Basque Research Foundation. The group also received funds from the breast cancer patient’s charity Katxalin and from Roche Farma S.A. JILV is funded by an AECC PhD Fellowship (PRDGI19007LOPE).

## Data availability statement

The data for this study are available upon reasonable request.

## Consent for publication and competing interests

We verify that each author consents the publication of this manuscript and authors have no conflicts of interest to declare.

## Notes

**Conflict of interest:** The authors have declared that no conflict of interest exists.

### Competing Interest Statement

The authors have declared no competing interest.

### Funding Statement

This work was funded by Spanish Ministry of Science and Innovation ISCIII (P20/01253, CP18/00076 and FI19/00193) and European Regional Development (FEDER) funds, Basque Department of Health (2020111040), Fundacion SEOM (SEOM Avon Fellowship 2020) and Ikerbasque Basque Research Foundation. The group also received funds from the breast cancer patients charity Katxalin and from Roche Farma S.A. JILV is funded by an AECC PhD Fellowship.

### Author Declarations

Ethics committee/IRB of Spain Health Authority and Diputacion de Gipuzkoa gave ethical approval for this work

### Summary of Updates

There are not significant differences between this version and the previous version uploaded. Only misprints and text/figures formats have been modified.

