## Supplemental Figures S1, S2 & S3 for "A prospective study on tumour response assessment methods after neoadjuvant endocrine therapy in early oestrogen receptor positive breast cancer"

**Supplementary material online**

Supplementary figure legends:

**Figure S1.** Changes in tumour levels of: Ki67 (A), Oestrogen receptor (B) and Progesterone receptor (C) before and after NET.

**Figure S2.** Association between tumour cellularity size (A), tumour cellularity (B) and pathological tumour size (C) with Ki67 levels at surgery. Spearman correlation coefficients (rho) and *p* values are shown.

**Figure S3**. Comparison of TCS quartiles (Q1, Q2, Q3 and Q4) with Ki67 levels at surgery (A) and ∆Ki67 (B). *p* values were calculated using Mann Whitney test.

### Supplementary Figure S1

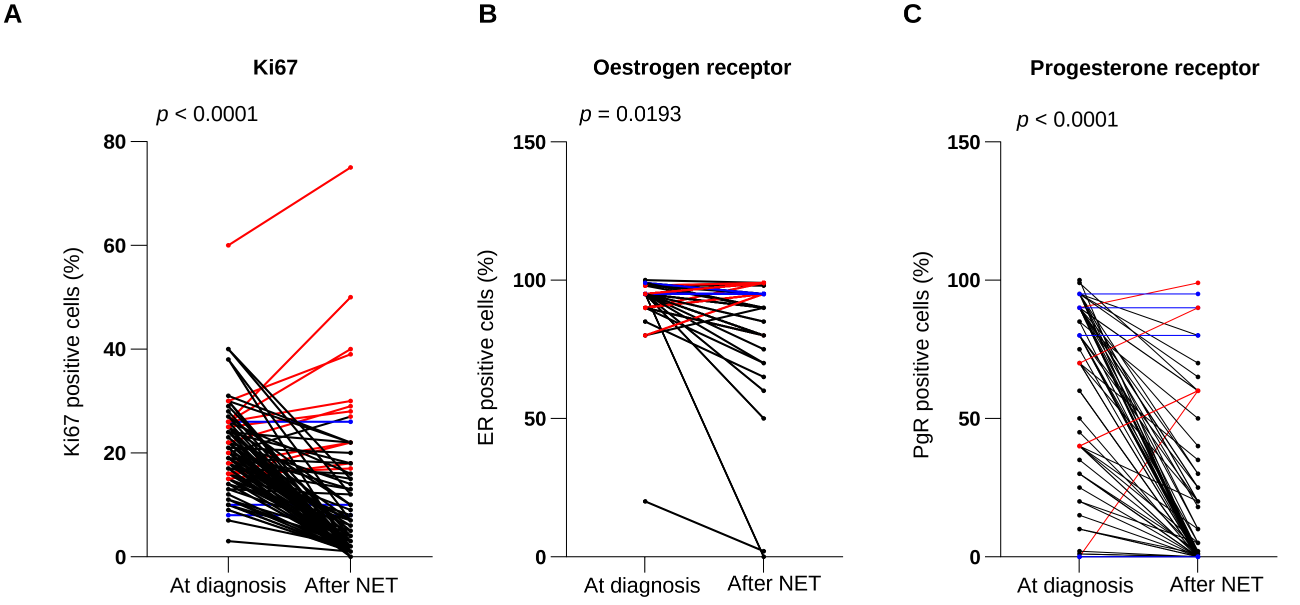


Positive cells (%): **mean** (range)

| | Ki67 | | ER | | PgR | |
|---|---|---|---|---|---|---|
| | At diagnosis | After NET | At diagnosis | After NET | At diagnosis | After NET |
| Positive cells (%): **mean** (range) | **20** (3-60) | **9** (0-75) | **94** (20-100) | **90** (0-99) | **63** (0-100) | **14** (0-99) |

*Change between at diagnosis and after NET:*

*Decrease; Increase; No change*

### Supplementary Figure S2

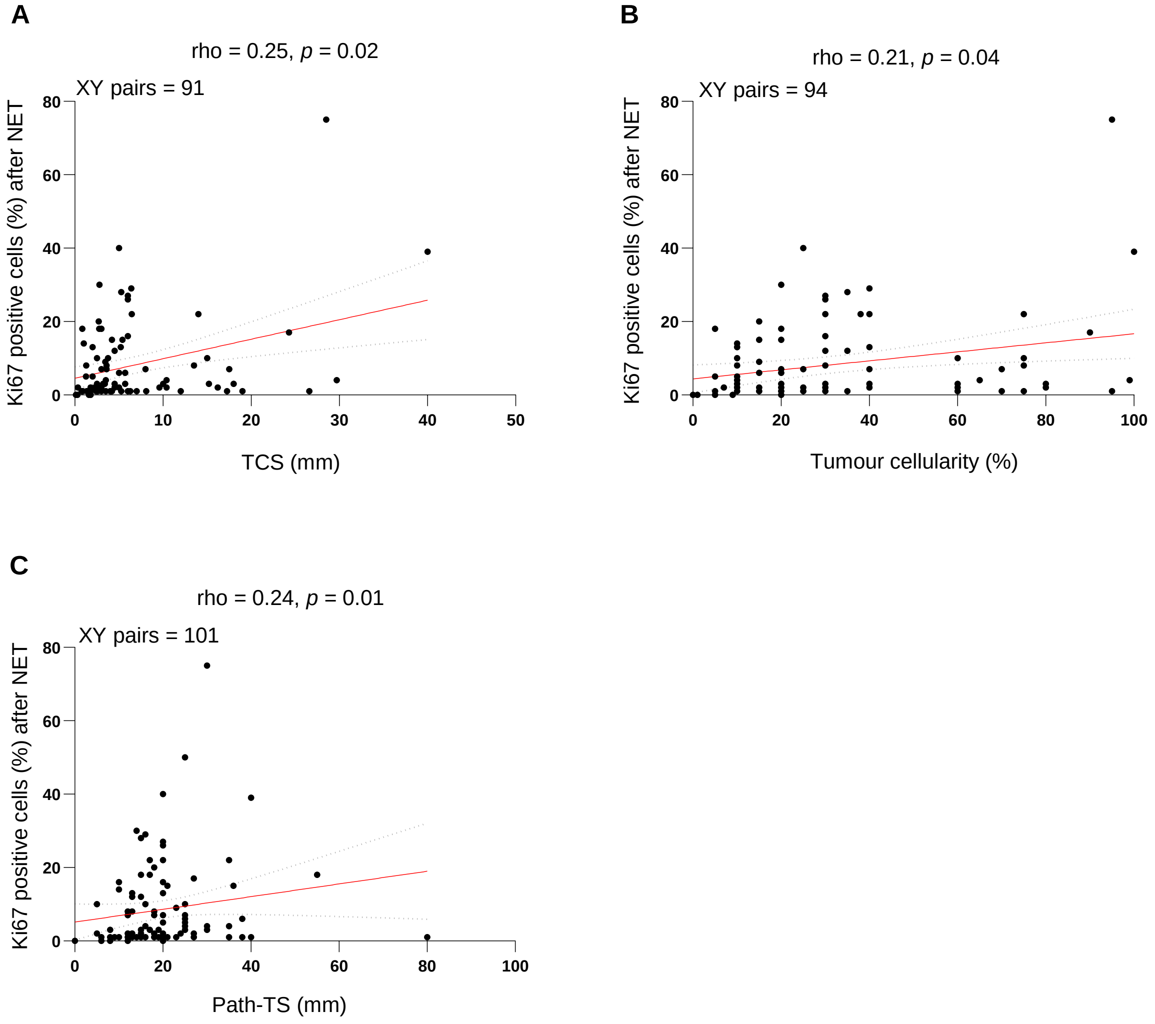

### Supplementary Figure S3

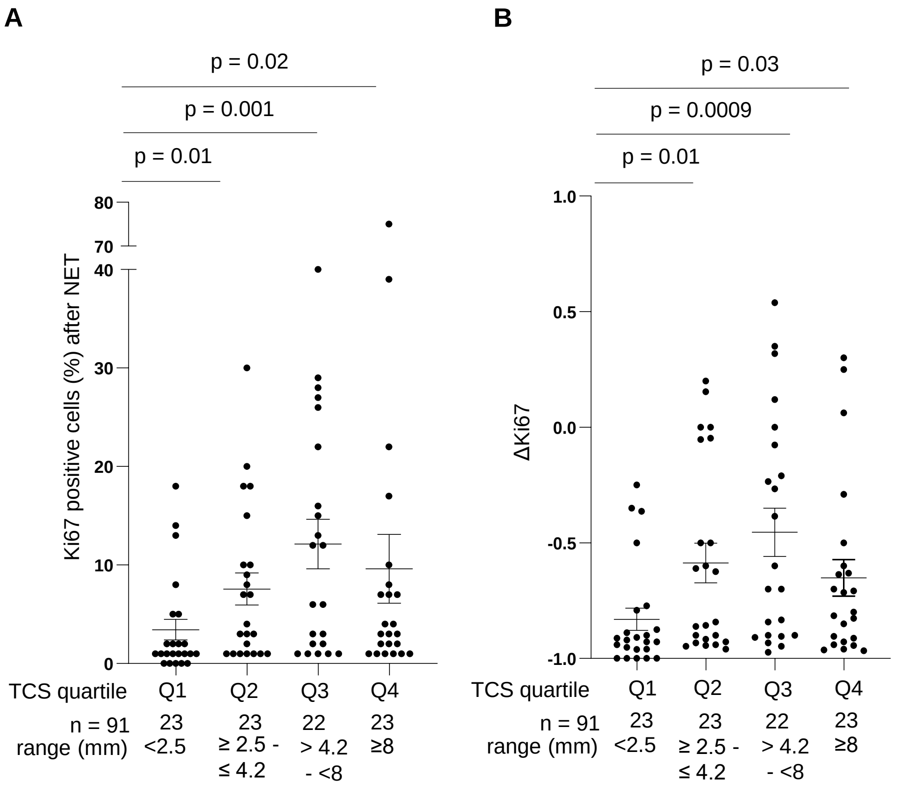
